# AI support for ethical decision-making around resuscitation: proceed with care

**DOI:** 10.1101/2020.08.17.20171769

**Authors:** Nikola Biller-Andorno, Andrea Ferrario, Susanne Joebges, Tanja Krones, Federico Massini, Phyllis Barth, Georgios Arampatzis, Michael Krauthammer

## Abstract

Artificial intelligence (AI) systems are increasingly being used in healthcare, thanks to the high level of performance that these systems have proven to deliver. So far, clinical applications have focused on diagnosis and on prediction of outcomes. It is less clear in what way AI can or should support complex clinical decisions that crucially depend on patient preferences. In this paper, we focus on the ethical questions arising from the design, development and deployment of AI systems to support decision-making around cardio-pulmonary resuscitation leading to the determination of a patient’s Do Not Attempt to Resuscitate (DNAR) status (also known as code status). The COVID-19 pandemic has made us keenly aware of the difficulties physicians encounter when they have to act quickly in stressful situations without knowing what their patient would have wanted. We discuss the results of an interview study conducted with healthcare professionals in a university hospital aimed at understanding the status quo of resuscitation decision processes while exploring a potential role for AI systems in decision-making around code status. Our data suggest that 1) current practices are fraught with challenges such as insufficient knowledge regarding patient preferences, time pressure and personal bias guiding care considerations and 2) there is considerable openness among clinicians to consider the use of AI-based decision support. We suggest a model for how AI can contribute to improve decision-making around resuscitation and propose a set of ethically relevant preconditions – conceptual, methodological and procedural – that need to be considered in further development and implementation efforts.

## AI-BASED DECISION SUPPORT IN HEALTHCARE

An artificial intelligence (AI) is a computer system performing tasks that normally require human-level intelligence, like the identification of patterns in data and the generation of predictions to assist decision-making processes. At the core of modern AI lies machine learning, which “transforms the inputs of an algorithm into outputs using statistical, data-driven rules that are automatically derived from a large set of examples, rather than being explicitly specified by humans” [1]. Deep learning is a form of machine learning that allows to automatically learn multiple layers of data representations, with minimal data preprocessing by humans [2]. Thanks to the increase in available data and computational power, machine and deep learning models can be trained on massive datasets to deliver high performance in tasks from multiple domains, including healthcare.

It has been recognized for some time that machine learning algorithms are not perfect. They can be subject to biases – caused, for instance, by incomplete training data sets or misclassification errors -, which may potentially lead to serious real-life implications such as amplifying socioeconomic disparities in healthcare [3]. In spite of these challenges, machine learning-powered AI systems already run in a myriad of healthcare applications, from diagnostics to prevention, drug discovery, treatment recommendations and operational excellence. For example, AI systems using deep learning have been successful in multiple medical imaging use cases [4-8]; they have proven to predict reliably the risk of imminent suicide attempts [9], and they have been used to assess the probability of patients developing serious conditions or being transferred to palliative care [10].

An increasing number of scientific contributions aim at the comparison of the performance of AI systems with the performance of human experts in the same healthcare domain: it has been shown [11] that AI systems using deep learning can match the diagnostic performance of healthcare professionals. In some medical domains, the use of AI systems will replace a considerable part of the work of human experts [12]. However, performance comparisons between AI systems and human experts suffer from the difficulty in reproducing and comparing results due to the lack of a unified approach [11,13,14]. Yet, it is reasonable to assume that AI systems will increasingly gain in epistemic authority, even though the assistive role of AI is frequently emphasized [15]. This renders a definition of ethical framework conditions for the use of AI-based decision support in healthcare all the more important and timely [16]. We have chosen to approach the issue through the study of a specific potential application, an AI-based support system for decisions around resuscitation.

## RESUSCITATION DECISIONS AS AN ADVANCED TEST CASE FOR AI-BASED DECISION SUPPORT?

Critical healthcare issues such as the cardiopulmonary resuscitation of patients require complex decisions with significant ethical implications [17]. Moreover, such decisions frequently need to be taken in relatively short time frames and under stressful conditions. The current pandemic, with its high number of people falling severely ill very quickly, has once more made us keenly aware of the difficulties physicians encounter when they have to act without having a chance to know what the patient would have wanted. Humans making such decisions must integrate multiple sources of information and calibrate them with personal, social and ethical standards. With AI-based systems becoming mature for clinical decision support, such information processing and decision-making can be assisted computationally. Algorithmic suggestions can be consistent with patient preferences and likely outcomes without getting compromised by stress, time pressure, personal bias, conflicts of interest and fear of legal consequences that may influence provider perspectives and the end-of-life conversations with their patients.

The status quo is less than perfect: A national audit in England showed that almost 40,000 patients every year are receiving DNAR orders without their consent or knowledge of their families^1^. Physicians seem to be making these decisions for a good part relying on their own judgment, based on medical parameters, experience and personal values [18]. Often no advance directive and no surrogate decision-maker are available, so physicians or a legal representative unfamiliar with the patient need to step in [19]. Patients often have insufficient access to professional advance care planning and a limited understanding of a life-threatening resuscitation situation, so it is difficult for them to assess the situation and define their resuscitation status ahead of time (e.g. in an advance directive). This means that a significant part of patients arrives in the hospital arrives without having made any indication at all of their will regarding resuscitation. Others may base their decision on inaccurate assumptions. For instance, cardio-pulmonary resuscitation is not as successful as it is often perceived [20], so patients may overestimate its benefits and disregard its potential harms when taking their decision. Also, it has been shown that relatives acting as surrogate decision makers on life-sustaining treatment frequently feel overburdened [21] and make choices that often do not resonate with the patient’s preferences [21,22].

A personalized, AI-based decision support system that is readily available to patients, relatives and healthcare professionals could make a significant contribution towards improving the status quo, in addition to other initiatives like advance care planning (ACP), that intend to improve decision-making by providing relevant information and a structured decision-making process to the patient [23]. Decision sciences are currently focusing on a variety of value clarification methods [24], and algorithmic support would be a novel approach helping people understand how others with whom they share certain features and values have decided in a similar situation [25]. In this sense, AI would act as a DNAR status decision support for (potential) patients (cf. the patient-centric application in Graph 1). AI-based decision support might also be helpful for legal representatives having to decide on behalf of incapacitated patients, ideally in an ACP by proxy process, without having a clear sense of what the patient would have wanted. It might also support healthcare professionals (HCPs) who need to make resuscitation decisions without the possibility to consult an advance directive or legal representatives, by helping clarify which option most likely corresponds to what the patient would have chosen (cf. the proxy/provider-centric application in Graph 1). Given the limitations of algorithmic decision-making (with a view to the risk of bias and to the dependency on suitable training data), the highly personal nature of the decision and its far-reaching consequences, the AI system would be conceived to play a consultative rather than a prescriptive role, it would not replace but support human decision-making.

Considering the use of AI to support decision-making around resuscitation quickly raises a large set of questions about feasibility, appropriateness and potential impact: Are suitable data sets available for an algorithm to train on, given that many human DNAR decisions today are taken in non-ideal circumstances? Should an AI system be modeled after patient preferences or outcomes, or both? How would users perceive such a system? For successfully implementing decision support, context factors matter greatly [26]: Would an AI system be a good fit with clinical routines? Would users consider its outputs interpretable and trustworthy? And how can we evaluate if an AI system does more good than harm?

## RESUSCITATION DECISION-MAKING IN THE HOSPITAL AND THE USE OF AI SYSTEMS: VIEWS OF HEALTHCARE PROFESSIONALS

We conducted a qualitative pilot study inviting healthcare professionals’ perspectives to probe the potential and limitations of an AI-based decision support around resuscitation. Although resuscitation decisions need to be taken across the spectrum of medical care, we chose to focus on a hospital setting, assuming that the majority of resuscitation attempts occur in the in-patient sector, rendering resuscitation status particularly important, and that digital documentation was most advanced. We chose a university hospital in the region for our exploration, for various reasons: 1) Code status is systematically collected from all patients and documented in the electronic hospital information system (since 2013), 2) The hospital has got a large number of documented resuscitation decisions (120’000 in total), spread across the different departments, 3) Unlike many other healthcare institutions, the hospital offers advance care planning (ACP) for patients by trained professionals including so-called physician orders for life-sustaining treatment (POLST, “an approach to end-of-life planning based on conversations between patients, loved ones and health care professionals”^2^) [27], so that the decisions resulting from this process can be considered fairly well informed and well reflected [28]. A significant part of healthcare providers also receive obligatory training on the ethical and procedural rules governing resuscitation [29], which are defined by national guidelines.

We therefore assumed that resuscitation or code status (we use the term interchangeably) was a topic that healthcare professionals were clearly aware of, and that it was part of everyday routine for many of them, so that interview partners could provide us with substantive answers. We were also particularly interested in this setting as a candidate for a potential later training site of an AI system. Several other studies have already probed factors influencing (human) resuscitation decisions in other hospitals in Country X [30-32], which provided a valuable general background for our work.

### Methods

In summer 2019, we approached physicians and nurses working in a Swiss university hospital with a request for an interview for a study on resuscitation practices. The sample was designed for maximum variation. We selected individuals to represent those clinical disciplines that we assumed would most frequently be confronted with resuscitation decisions and that at the same time covered a good part of the in-hospital pathway (emergency care, internal medicine, intensive care, surgery, palliative care). All study participants had clinical experience but differed in level of seniority.

Participation in the study was voluntary; all participants were contacted by e-mail, and interviews took place at the hospital. Ethics approval was obtained by the relevant IRB in accordance with legal provisions. All participants gave informed consent and the interviews (in German) were recorded on tape. We conducted 11 semi-structured interviews (6 female, 5 male, 7 physicians, 4 nurses) of 25 to 45 minutes, which took place at the university hospital. They were audio-recorded, transcribed and analyzed following standards for expert interview analysis [33].

The guide was developed based on an international literature review of papers on resuscitation status decision handling, the historical development of patient’s involvement regarding resuscitation status, case studies about controversial handling of a patient’s resuscitation status, and ethical guidelines regarding DNAR orders. We complemented it with a specific information research for the University Hospital Zurich (patient information brochure regarding resuscitation status, e-learning program about resuscitation status discussion with the patient). Questions covered current practices regarding determination and documentation of the decision to attempt resuscitation (DNAR) or not, the decision-making process, perceived challenges and reactions to the possibility of AI-based support. The interview guide^3^ was constructed to 1) to highlight specificities and procedural criticalities in the existing processes around resuscitation for different units at the hospital, with a focus on the update of resuscitation status entries, and 2) to discuss with healthcare professionals the possibility of using AI systems to support their decision-making on resuscitation.

### Results

Our study aimed to provide insights into resuscitation status decision-making in a large university hospital of international renown that might serve as a future site for training and pilot implementing a prototype of an AI-based decision support system. We wanted to understand where the issue of resuscitation status became relevant over the course of patient pathways and what the challenges were.

The aspirational goal for the studied institution is as clear as it is ambitious: The resuscitation status of all patients has to be established based on patient preferences – determined either in conversation with the patient, through an advance directive or a legal representative – and needs to be respected unless there are overriding considerations, such as clear cases of inappropriate or even futile care. An exception regards newly admitted patients whose condition and urgent medical needs do not allow for a determination of their resuscitation preferences [34]. Code status has to be continuously updated and documented in a way that allows easy access to the respective treatment team.

Based on the insights gleaned from the interviews, the graph shows where resuscitation status becomes relevant over patient pathways, highlighting and situating key challenges.

We identified several core challenges regarding the institutional goal of documenting up-to-date and patient preference-based resuscitation status information of all in-patients, which emerged from our interviews with HCPs (cf. Box 1)

Interview partners demonstrated clear awareness of the importance of code status documentation and its challenges. When the issue of AI came up in the interviews, there was a general openness towards considering AI-based decision support to help improve the status quo. At the same time, interviewees volunteered considerations – very much a brainstorming, as this potential application of AI is still a highly innovative idea – regarding the role, strengths and limitations of such a system if used in routine hospital care (cf. Box 2).

## AI SUPPORT FOR DNAR DECISION-MAKING IN THE HOSPITAL: TOWARDS A FRAMEWORK OF PRECONDITIONS FOR ETHICAL USE

Our study shows that in a highly functional university hospital setting there is ample room for improving the current system of determining and documenting patients’ code status (Box 1). HCPs are open towards considering AI-based support, but rightly point to the need to clarify issues such as the role of AI in relationship to human decision-making (Box 2). In addition, legal issues regarding, for instance, liability in case of errors will have to be addressed. We do think AI carries high potential for improving resuscitation-related decision-making, given the status quo. However, we are aware that the development of such a system is ethically demanding and requires attention to a set of preconditions as a first input into what we hope will grow into a broader debate.

### Conceptual preconditions

The AI-based system is an assistive advice, to be used either by patients reflecting on the choice of their DNAR status or by proxies and/or physicians deliberating if resuscitation would reflect an incapacitated patient’s will (cf. Graph 1). It does not have decisional authority and should never replace conversations with the patient, legal representatives or within the treatment team. The system can act as a conversation prompter, tie breaker or second opinion; it may invite self-critical reflection of the physician in charge or possibly of relatives and even patients. It may act as a support tool in case no information about a patient’s will is available.

The AI-based system predicts which resuscitation status a patient would have chosen after well-informed considerations of the benefits and harms (as presented in state-of-art, evidence-based decision aids). AI-based algorithms, given appropriate training data, could also predict under what conditions (if any) this choice would change, for instance when the likelihood for survival drops below a certain value. In this scenario, the outcome-based preference predictions would then be compared with the likely outcomes (possibly also predicted by an algorithm) for an individual patient, and the code status adjusted accordingly.

When training algorithms, the use of more easily accessible but unsuitable surrogate parameters for well-considered patient choice – such as patients having received CPR in a hospital or having been attributed a certain code status, should be avoided as they may not reflect patients’ wishes. They may perpetuate or even reinforce the flaws of the current status quo of DNAR decision-making at the site(s) from which training data are collected.

### Methodological preconditions

Data quality needs to be assessed to reduce the risk of “garbage in, garbage out”. Patient resuscitation preferences used for training the AI system (the “ground truth”) need to be elicited in a state-of-the-art way, e.g. through ACP conversations. A simple model would conceive of patient preferences as categorical (e.g., no CPR under any circumstances). The machine-learning task is therefore a prediction of a dichotomous outcome (DNAR yes / no), given available patient features, which are recorded in the EHR. The decision to opt for or against CPR in the case of cardiac arrest can be assumed to depend on a number of factors, among them a patient’s health status, life expectancy, current quality of life, perceived social obligations towards others, religious beliefs or deeply held secular values. Good training data will include such factors, which need to be encoded in health records, through discussions with the patient, relatives or other means. The algorithm will then be designed to predict the DNAR status based on these features, for example by applying a similarity metric to determine what “similar patients” would decide in the same circumstances [35]. Ideally, decisions taken on the basis of these preferences can be validated ex-post by physicians, relatives or even patients.

It is theoretically possible to conceive an outcome-sensitive code status prediction algorithm. Here, ACP conversations would result in several resuscitation status/outcome tuples. Each tuple reflects the DNAR choice given a possible patient outcome (after CPR), such as chances for survival to discharge do not drop below a certain value or the risk for severe cognitive deficits does not exceed a certain percentage. The AI prediction would then first determine the likely outcome of the patient (after CPR) and would then proceed as above and determine DNAR status given the likely outcome.

The development of an AI-based resuscitation decision assistant should make efforts towards explainability [36,37] and include elements of explanations from social sciences [38]. An opaque (i.e. “black box”) system, even if performing well, may not find acceptance and be trusted to advice on matters of life or death; issues of user’s trust, accountability, liability and data protection have to be addressed in accordance with existing and emerging standards [39-41]. The system has to be monitored for any bias and to ensure the quality and fairness of its advice.

### Procedural preconditions

The embedding of the AI system in healthcare processes will be decisive for its performance and user acceptance. Compatibility with the hospital information system and access management are relevant design features. User training is key: It has to cover the technical aspects of the AI system, and provide a view on how to integrate its predictions into clinical decision-making and into the communication with patients, relatives and staff in accordance with best practice principles of shared decision-making [42,43]. It should also aim at improving communication skills of the AI system users and raise awareness on the risks and limitations of AI support in decision-making [44]. This should allow countering any tendencies to avoid difficult conversations with patients or family members about DNAR status by resorting to algorithmic predictions, as well as minimizing the likelihood to outsource a core clinical task – establishing a patient’s DNAR status – and its potential implications for the patient-provider- relationship to an AI system. We sketch examples for potential use cases in Box 3.

The examples given in Box 3 hints at the many questions that the implementation of such a system would raise: What information can or should the system reveal about how it calculated the preferences (transparency, explainability)? How do we know an AI system is good enough for use in clinical routine? What if the AI system yields a result the physician considers highly implausible or inappropriate? etc.

Deploying an AI-based support for resuscitation decisions is a high risk, high reward undertaking. Whereas the potential to improve decision-making and alleviate burden on treatment teams is considerable, particularly when little is known about a patient and when various courses of action are justifiable, a poorly designed system can reinforce current flaws in decision-making and introduce new ones. Our qualitative pilot provided just a first exploration of health professionals as one potential user group. Critical scrutiny from many perspective – including, importantly, those of patients and legal representatives – will be key to increasing chances for success, not only with a view to user acceptance but also to responsible use of AI.

## CONCLUSION AND OUTLOOK

Leveraging AI for healthcare decision support raises critical questions at the interface of medicine, ethics and computer science. These questions include issues of bias and fairness as well as of autonomy and accountability [45-47]. The exploration of an example for a specific decision support system provides an opportunity to discuss and address ethical and policy concerns in a very concrete, hands-on way [41]. A carefully reflected, well-designed AI-based system can have an immediate, significant and practical impact to personalized healthcare by contributing to better outcomes of critical healthcare decisions. However, it is of prime importance that framework conditions are defined such that they justify citizens’ and health professionals’ trust in the AI system. Evaluation standards for the performance of AI-based decision-support systems will be urgently needed to make sure systems can be discarded if unsuitable or improved.

Once AI systems are unequivocally recognized as being more accurate and reliable than human practitioners in generating predictions, or suggesting treatments and diagnosis, a shift in epistemic authority may occur [13]. This shift will raise questions about a potential obligation to rely on these systems when engaged in medical decision-making processes [48-50] – an important debate that is, however, well beyond the scope of this paper.

## Data Availability

The interview guide has been translated into English and is available from the authors upon request. We also produced English interview summaries and translations of key interview quotes.

## Acknowledgments

We gratefully acknowledge our interview partners who generously supported our study with their time and insights. We would also like to thank Prof. Dr. David Blum and Dr. Settimio Monteverde for valuable comments and advice.

## Footnotes

1 https://www.telegraph.co.uk/news/2016/05/01/unforgivable-failings-in-end-of-life-care-revealed-40000-dying-p/

2 www.polst.org, last accessed on 25 July 2020.

3 The interview guide has been translated into English and is available from the authors upon request. We also produced English interview summaries and translations of key interview quotes.

## Notes

### Competing Interest Statement

The authors have declared no competing interest.

### Funding Statement

This study formed part of a larger fellowship project on Digital support of decision-making in health care (led by Nikola Biller-Andorno) at the Collegium Helveticum, an Institute of Advanced Studies carried by the University of Zurich, the Swiss Federal Institute of Technology (ETH) Zurich and the Zurich University of the Arts (ZHdK).

